## Supplemental data for "Defective CAPSL function causes impaired retinal angiogenesis through the MYC axis and is associated with familial exudative vitreoretinopathy"

Figure S1-S8, Table S1 to S3.

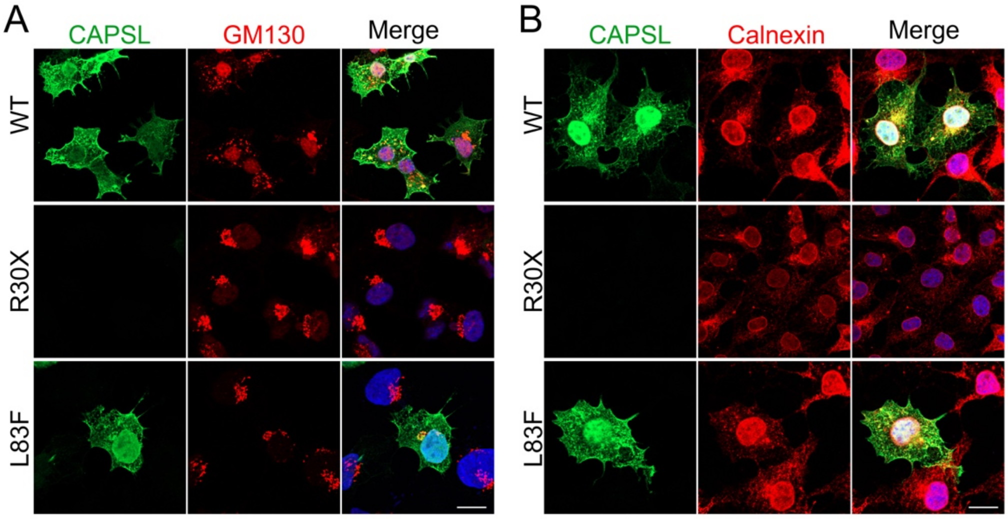


**Figure S1. Subcellular localization of CAPSL variant proteins.**

(A) Transfected 293T cells with wild-type CAPSL plasmid and variant plasmids were fixed and stained for CAPSL (green), GM130 (red, Golgi), and DAPI (blue, nucleus). Scale bar: 5 μm. (B) Transfected 293T cells with wild-type CAPSL plasmid and variant plasmids were stained for CAPSL (green), Calnexin (red, Endoplasmic Reticulum). Scale bar: 5 μm.

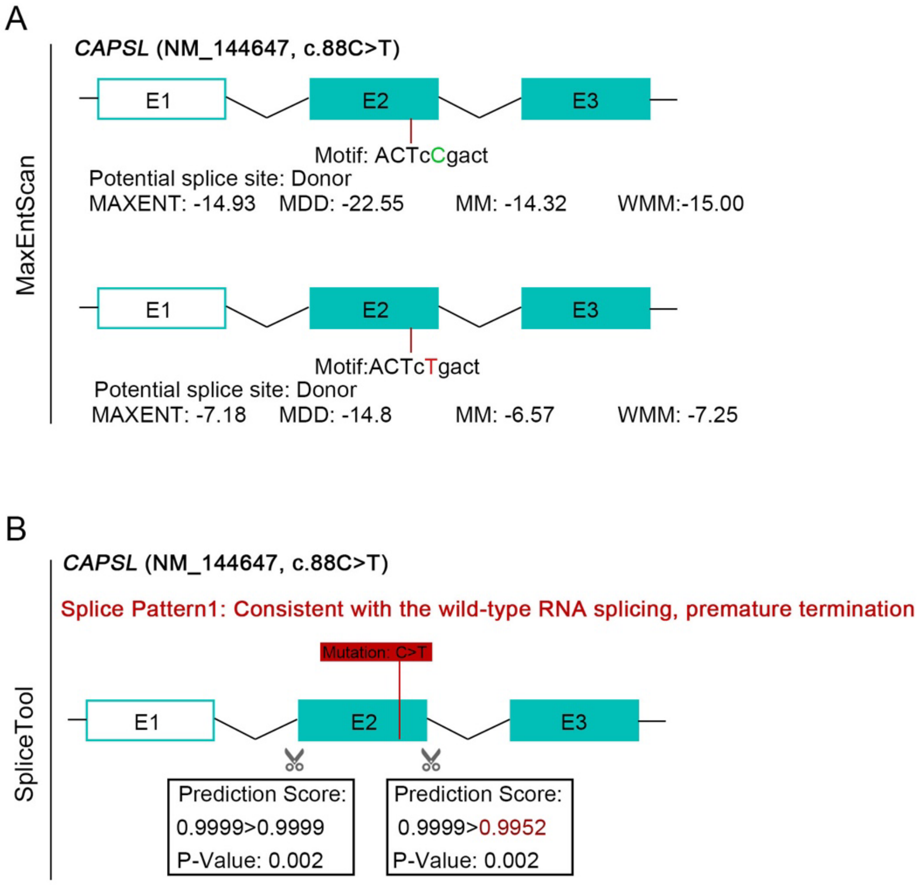


**Figure S2. Bioinformatic prediction of an impact on splicing of the pathogenic variant c.88C>T in *CAPSL*.**

(A) MaxEntScan was used to investigate the impact of the variant on the formation of a cryptic donor splice site. Four different models were used: MAXENT: Maximum Entropy Model; MDD: Maximum Dependence Decomposition; MM: First order Markov Model and WMM: Weight Matrix Model. (B) SpliceTool predicts an impact on splicing for the variant c.88C>T.

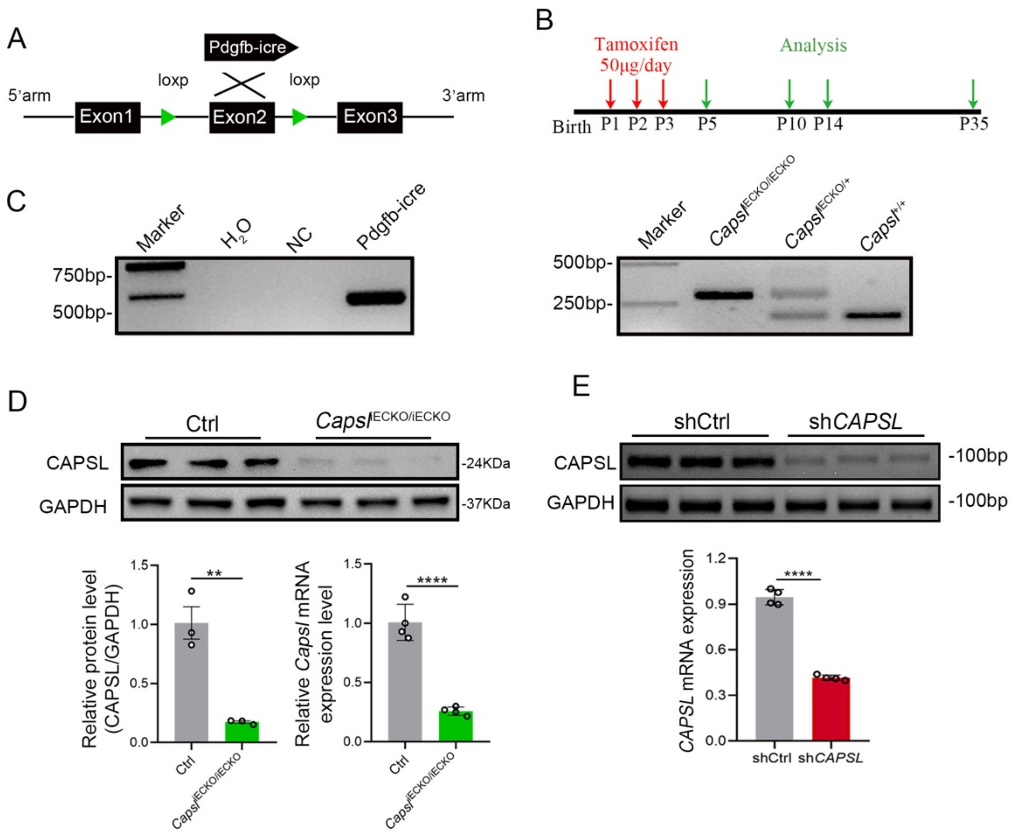


**Figure S3. Construction of EC-specific *Capsl* knockout mice model and CAPSL knockdown HRECs cell model.**

**(A)**Strategy to generate a conditional *Capsl* allele in which exon 2 are knockout specifically in endothelial cells. **(B)** Diagram depicting the tamoxifen injection and experiment schedule for EC-specific deletion of *Capsl* in retinal vessels from P1 to P3 and their analyses timeline. **(C)** PCR of genomic DNA from Ctrl and *Capsl*$^{iECKO/iECKO}$ pups. Recombination of the floxed *Capsl* allel occurs only in Tamoxifen-injected animals that are Pdgfb-iCreER-positive. **(D)** Immunoblot and quantification of CAPSL knockout efficiency at both protein and mRNA level. Error bars indicate the SD. **P < 0.01, ****P < 0.0001, by Student's t test (n = 3). **(E)** RT-PCR and RT-qPCR analysis of knockdown efficiency of lentivirus carrying sh*CAPSL*. Error bars indicate the SD. ****P < 0.0001, by Student's t test (n = 3).

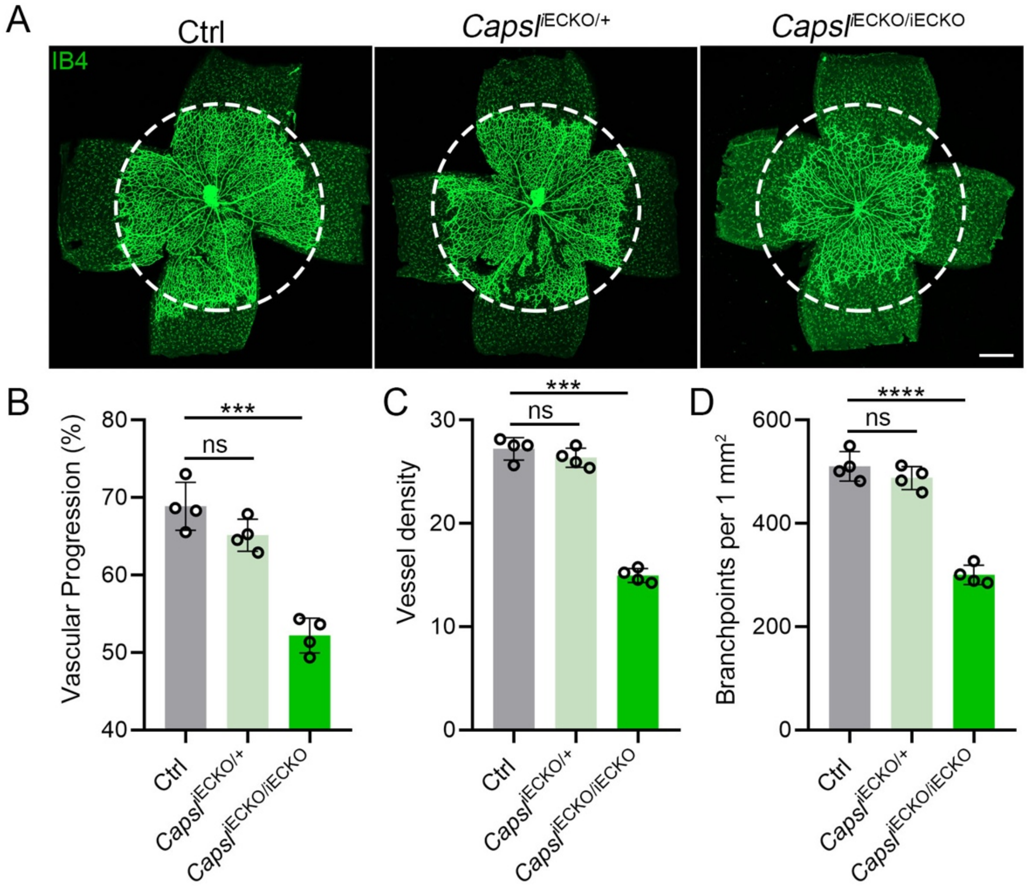


**Figure S4.** ***Capsl*** **heterozygous mice exhibited no significant defect in angiogenesis.**

**(A)** Flat-mounted retinas obtained from P5 *Capsl*$^{iECKO/+}$, *Capsl*$^{iECKO/iECKO}$, and littermate Ctrl mice were stained with Isolectin-B4 (IB4) to visualize blood vessel. Dashed circle mark3 the edge of the developing retina vessel in Ctrl mice. Scale bar: 250 µm. **(B-D)** Quantification of retinal vascular development parameters, including vascular progression, vessel density, and branchpoints. Error bars indicate the SD. ***P < 0.001, ****P < 0.0001, by Student's t test (n = 4).

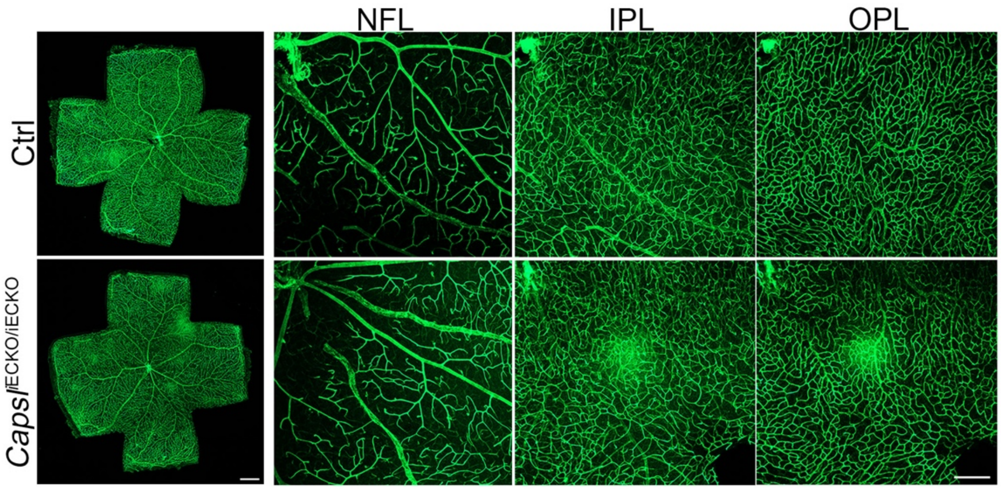


**Figure S5.** ***Capsl*** **deletion does not affect vessel maturation.**

Flat-mounted retinas stained with Isolectin-B4 (IB4) at P21 Ctrl and *Capsl*$^{iECKO/iECKO}$. Optical sections of z-stacked confocal images were divided to represent the nerve fiber layer (NFL), inner plexiform layer (INL), and outer plexiform layer (ONL). Scale bar: 250 μm (left panel) and 100 μm (right panel)

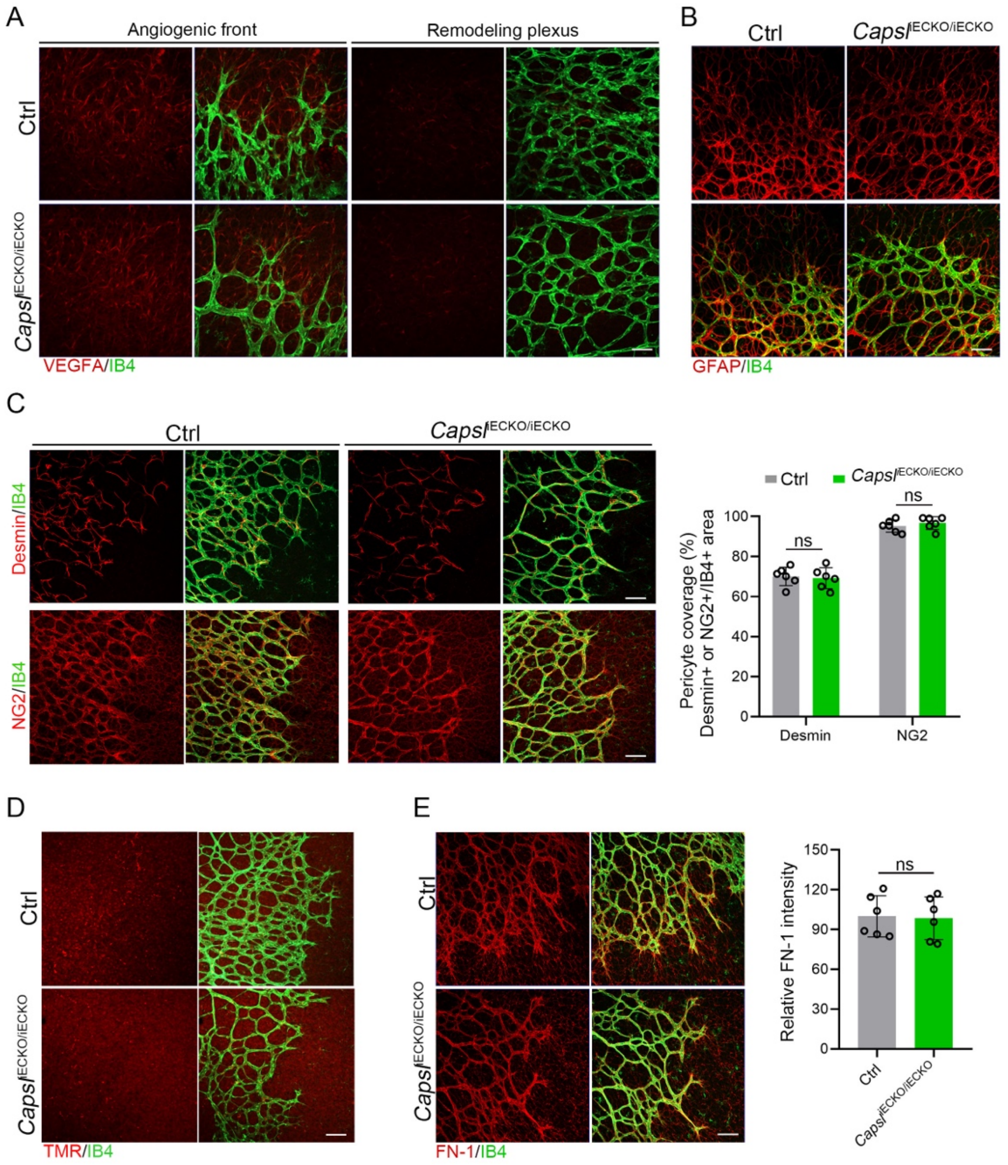


**Figure S6. Normal state of other cell types and ECM deposition in *Capsl* knockout mice retina.**

**(A)** Co-staining of VEGFA (red) and IB4 (green) at angiogenic front and remodeling plexus in P5 retinas of Ctrl and *Capsl*$^{iECKO/iECKO}$ mice. Scale bar: 50 μm. **(B)** Co-staining of Collagen IV (red) and IB4 (green) in P5 retinas of Ctrl and *Capsl*$^{iECKO/iECKO}$ mice. Scale bar: 50 μm. **(C)** Maximum intensity projections and quantification of pericyte coverage of the superficial vascular plexus in P5 retinas of Ctrl and *Capsl*$^{iECKO/iECKO}$ mice stained with Desmin/NG2 (red) and IB4 (green). Scale bar: 50 μm. Error bars indicate the SD. ns: no significance, by Student's t test (n = 6).

**(D)** Representative confocal images of biocytin TMR (red) and IB4 (green) in the retinal flat mounts from P5 Ctrl and *Capsl*$^{iECKO/iECKO}$ mice. Scale bar: 50 μm. **(E)** Immunofluorescence staining and quantitative analysis of FN-1 (red) stained ECM deposition in retinas form P5 Ctrl and *Capsl*$^{iECKO/iECKO}$ mice. Scale bar: 50 μm. Error bars indicate the SD. ns: no significance, by Student's t test (n = 6).

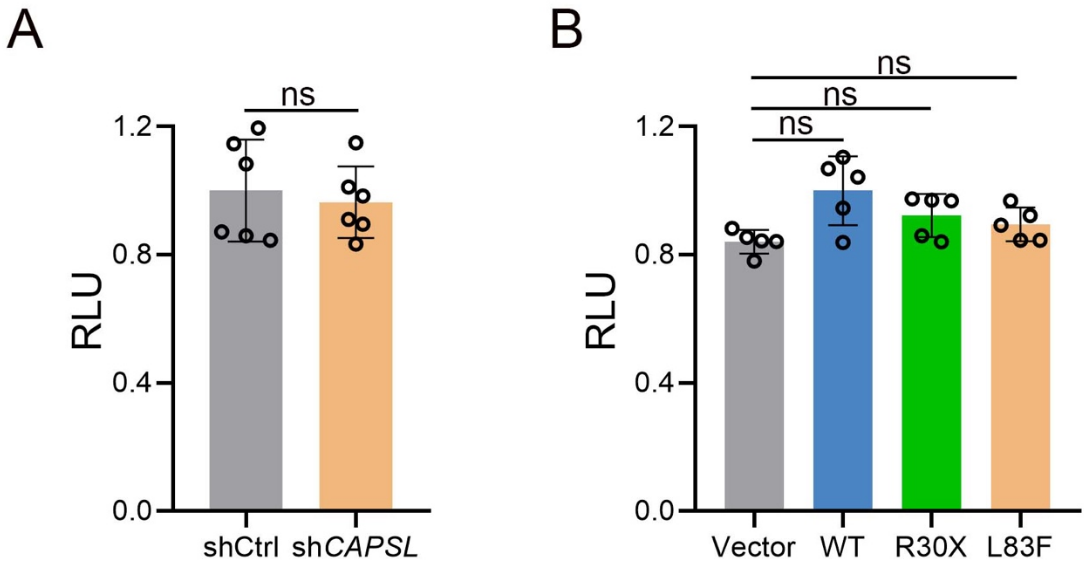


**Figure S7. CAPSL is not involve in Norrin/β-catenin signaling pathway.**

**(A)** Results of luciferase reporter assay of shCtrl-293STF cells and sh*CAPSL*-293STF cells. Error bars indicate the SD. ns: no significance, by Student's t test (n = 6). **(B)** Results of luciferase reporter assay of sh*CAPSL*-293STF cells were transfected with plasmids containing *CAPSL* (WT, R30X, or L83F) or empty vector. Error bars indicate the SD. ns: no significance, by Student's t test (n = 5).

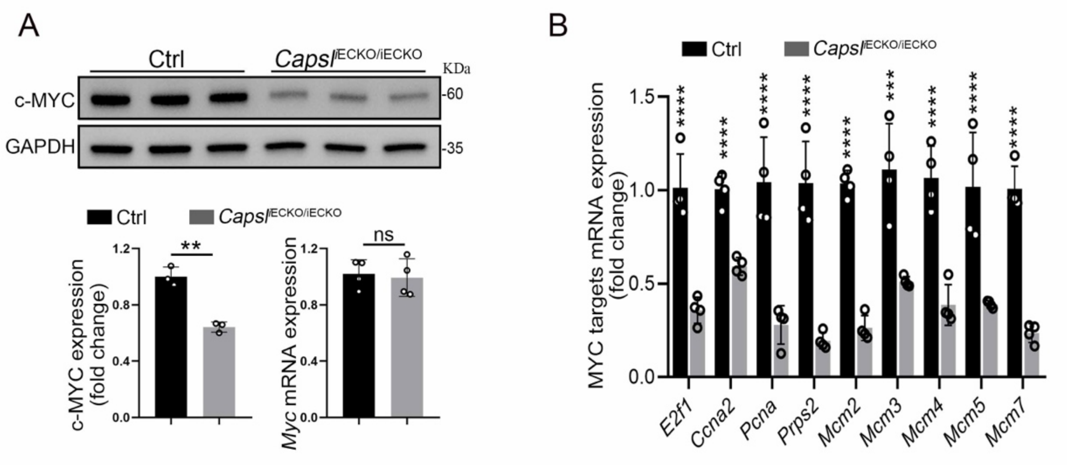


**Figure S8. *Capsl* depletion resulted in downregulation of MYC targets.**

(A) c-MYC expression level of P40 Ctrl and *Capsl*$^{iECKO/iECKO}$ mice lung tissue was quantified by western blot and RT-qPCR. Error bars indicate the SD. **P < 0.01, ns: no significance by Student's t test (n = 3). (B) Relative mRNA expression of MYC targets of P35 Ctrl and *Capsl*$^{iECKO/iECKO}$ mice lung tissue. Error bars indicate the SD. ***P < 0.001, ****P < 0.0001, by Student's t test (n = 3)

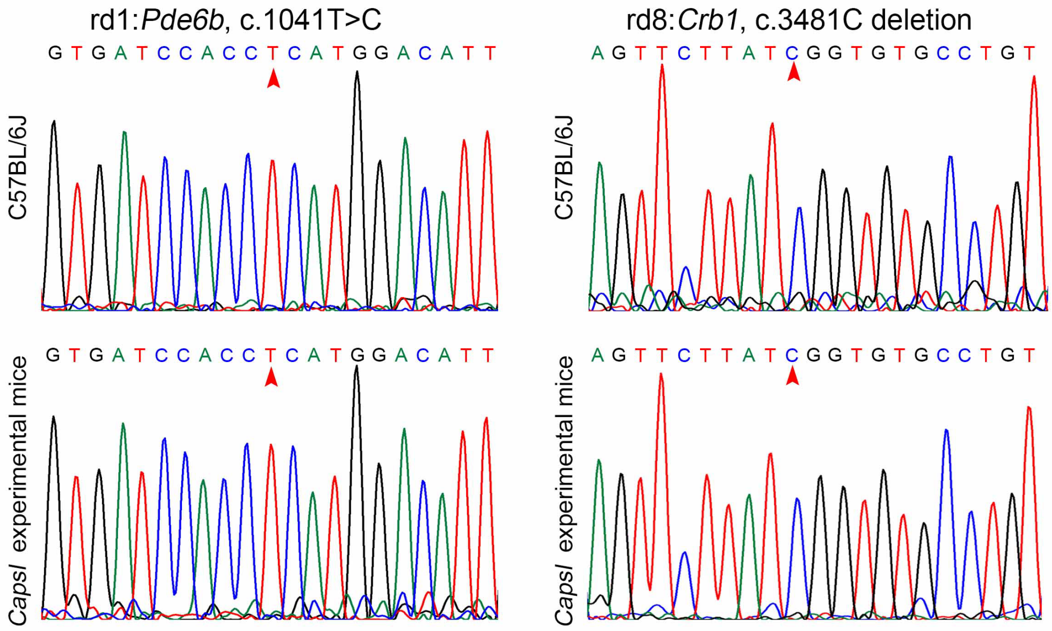


**Figure S9. Experimental mice do no carry confounding rd1 and rd8 mutations.**

(A) Sanger sequencing was performed to verify the *Pde6b* c.1041T>C in rd1 mice and experimental mice. **(B)** Sanger sequencing was performed to verify the *Crb1* c.3481C deletion in rd8 mice and experimental mice.

**Table S1. Pathological prediction of CAPSL variants**

| Software | Variant | Prediction |
|---|---|---|
| Mutation Taster | R30X | disease causing:<br>amino acid sequence changed<br>protein features (might be) affected<br>splice site changes |
| | L83F | disease causing:<br>amino acid sequence changed<br>protein features (might be) affected |
| Polyphen-2 | R30X | - |
| | L83F | Possibly damaging with a score of 0.777 (sensitivity: 0.85; specificity: 0.92) |
| PROVEAN | R30X | Score:-13.38, prediction: deleterious |
| | L83F | Score:-2.51, prediction: deleterious |

**Table S2. Antibodies used for immunofluorescence studies.**

| Antibody | Manufacturer | States | Cat. No. | Dilution |
|---|---|---|---|---|
| Isolectin-B4-Alexa488 | Thermo Fisher | USA | I21411 | 1:200 |
| VEGFA | R&D System | USA | AF-493-NA | 1:100 |
| Collagen IV | Millipore Sigma | USA | AB756P | 1:100 |
| Fibronectin-1 | Sigma | USA | F3648 | 1:100 |
| NG2 | Millipore Sigma | USA | AB5320 | 1:200 |
| Desmin | Abcam | UK | AB15200 | 1:100 |
| ERG | Abcam | UK | Ab92513 | 1:200 |
| GFAP | Cell Signaling Technology | USA | 80788 | 1:200 |
| DAPI | Cell Signaling Technology | USA | 4083 | 1:2000 |
| Donkey anti-Rabbit Alexa594 | Invitrogen | USA | A-21207 | 1:500 |
| Donkey anti-Goat Alexa594 | Invitrogen | USA | A-11058 | 1:500 |
| Donkey anti-Rabbit Alexa647 | Invitrogen | USA | A-31573 | 1:500 |

**Table S3. Primer sets used for RT-qPCR.**

| Name | Sequence 5'-3' | |
|---|---|---|
| *Capsl*-Mus | Forward | GGACTTGGCAGAGTGTTTCG |
| | Reverse | TCTGGCTCTGGACATTGGAG |
| *Myc*-Mus | Forward | TCAGACACGGAGGAAAACGA |
| | Reverse | CGTCTGCTTGAATGGACAGG |
| *Gapdh*-Mus | Forward | TGTGTCCGTCGTGGATCTGA |
| | Reverse | TTGCTGTTGAAGTCGCAGGAG |
| *CAPSL*-Homo | Forward | GCCAAGAAAAAGCTCACCAC |
| | Reverse | CTTTTTCCATGACCACAGCA |
| *MYC*-Homo | Forward | AACACACAACGTCTTGGAGC |
| | Reverse | GCACAAGAGTTCCGTAGCTG |
| *GFP*-Homo | Forward | ACGTAAACGGCCACAAGTTC |
| | Reverse | AAGTCGTGCTGCTTCATGTG |
| *GAPDH*-Homo | Forward | CCATGGGTGGAATCATATTGGA |
| | Reverse | TCAACGGATTTGGTCGTATTGG |

**Table S4. Antibodies used for western blot analysis.**

| Antibody | Manufacturer | States | Cat. No. | Dilution |
|---|---|---|---|---|
| CAPSL | Proteintech | China | 17174-1-AP | 1:2000 |
| MCM2 | Cell Signaling Technology | USA | 3619 | 1:1000 |
| MCM3 | Cell Signaling Technology | USA | 4012 | 1:1000 |
| MCM4 | Cell Signaling Technology | USA | 3228 | 1:1000 |
| MCM5 | Abcam | UK | AB76023 | 1:2000 |
| MCM7 | Cell Signaling Technology | USA | 3735 | 1:1000 |
| E2F1 | Abcam | UK | Ab92513 | 1:2000 |
| CyclinD1 | Cell Signaling Technology | USA | 55506 | 1:2000 |
| CyclinD2 | Cell Signaling Technology | USA | 3741 | 1:2000 |
| CyclinD3 | Invitrogen | USA | AB_2809647 | 1:1000 |
| CyclinE1 | Cell Signaling Technology | USA | 28808 | 1:1000 |
| CDK4 | Cell Signaling Technology | USA | 127903 | 1:1000 |
| PCNA | ABclonal | China | A12427 | 1:1000 |
| CDC42 | Proteintech | China | 0155-1-AP | 1:1000 |
| RHOA | ABclonal | China | A4855 | 1:1000 |
| Rac1 | ABclonal | China | A7720 | 1:1000 |
| MYL9 | ABclonal | China | A8738 | 1:1000 |
| P-MYL9-S19 | ABclonal | China | AP9055 | 1:1000 |
| GAPDH | Proteintech | China | 1E6D9 | 1:5000 |
